# Evaluation of Deep Learning Based Early Warning Indicators of Epidemic Outbreaks

**DOI:** 10.64898/2026.09.22.26363696

**Authors:** Burak Ayyorgun, Madhav Marathe, Aniruddha Adiga

## Abstract

In many natural dynamical systems, tipping points emerge where slowly changing external factors induce shifts from one state to another, known as critical transitions or bifurcations. In epidemic systems, detecting these transitions is important for outbreak preparedness. Various statistical indicators like lag-1 autocorrelation and variance have been employed as early warning signals (EWS) of critical transitions by showing how the dynamics of the system exhibit critical slowing down (CSD) near the transition. More recently, deep learning-based algorithms have been developed that attempt to more accurately detect and classify bifurcations; however, they are limited by the systems simulated in the training dataset, the length of the time series, and have not been rigorously evaluated on real-world epidemic data spanning multiple geographic regions. In this paper, we evaluate existing deep learning-based bifurcation prediction models, on a CDC-curated dataset containing weekly influenza hospital admissions across all fifty U.S. states and the nation as a whole. We develop a pipeline for data preparation, reproduction number (*R_t_*) estimation using the EpyEstim package (a Python implementation of the EpiEstim framework [4]), and bifurcation point identification. We systematically evaluate the models under several windowing strategies, including rolling, expanding, and fixed-length windows, and report accuracy, precision, recall, F1 score, and specificity. Our best-performing configuration (expanding window, length 100) achieves an accuracy of 90.09%, precision of 72.86%, recall of 96.23%, and F1 score of 82.93%. We additionally discuss Markov Regime Switching models as an alternative framework for detecting epidemic transitions. Our findings indicate that these models can generalize to real-world, state-level influenza data and are capable of online/real-time prediction.

## 1 Introduction

In many natural dynamical systems, tipping points emerge where slowly changing external factors induce shifts from one state to another, known as critical transitions or bifurcations. In epidemic systems, detection and classification of these critical transitions has shown to be important in preparing for outbreaks. A disease goes extinct if *R*_0_ < 1 and persists if *R*_0_ > 1, and a transcritical bifurcation occurs when *R*_0_ = 1, where both equilibria meet and exchange their stability.

Various statistical indicators like lag-1 autocorrelation and variance have been employed as early warning signals (EWS) of these critical transitions by showing how the dynamics of the system exhibit critical slowing down (CSD) near the transition. CSD occurs as a system approaches a bifurcation, where recovery from perturbations becomes increasingly delayed. While these statistical indicators can mostly identify bifurcations, they usually cannot predict the future state of the system and in addition, cannot classify the type of bifurcation (i.e. fold vs. transcritical vs. Hopf) [1].

More recently, deep learning-based algorithms have been developed that attempt to more accurately detect and classify bifurcations. However, the models developed by Bury et al. [1] are not effective at detecting bifurcations in real-world epidemic data. And the models developed by Chakraborty et al. [2] and Miry et al. [3] are not accurate on a state-by-state basis, where there are varying levels of noise and other environmental and/or demographic factors.

In this paper, we evaluate these pre-trained deep learning models on a CDC-curated dataset of weekly influenza hospital admissions across all fifty U.S. states and the nation as a whole. We develop a pipeline for data preparation and *R_t_* estimation, systematically assess model performance under different windowing strategies, and evaluate whether these models are suitable for real-time (online) bifurcation prediction. We also discuss the possible application of Markov Regime Switching models as an alternative to detecting bifurcations.

## 2 Prior Literature

### 2.1 Classical Early Warning Signals

Various statistical indicators have been employed as early warning signals (EWS) of critical transitions. These rely on the phenomenon of critical slowing down (CSD): as a system approaches a bifurcation, recovery from small perturbations becomes increasingly delayed, which manifests as rising variance and lag-1 autocorrelation in observed time series. While these statistical indicators can mostly identify approaching bifurcations, they usually cannot predict the future state of the system and cannot classify the type of bifurcation [1].

### 2.2 Bury et al. (2021)

Bury et al. [1] developed a deep learning algorithm using a CNN-LSTM architecture, trained on simulated time series from a variety of mathematical models exhibiting fold, Hopf, and transcritical bifurcations. The algorithm exploits information about normal forms and scaling behavior of dynamics near tipping points that are common to many dynamical systems, and was shown to substantially outperform traditional EWS indicators on both simulated and several empirical datasets. However, the models are not effective at detecting bifurcations in real-world epidemic data,the training data were based on generic mathematical models rather than epidemic-specific systems, and the models were trained only on time series detrended using a Lowess filter with span 0.20 [6].

### 2.3 Chakraborty, Miry et al. (2024–2025)

To address this, Chakraborty et al. [2] and Miry et al. [3] developed models trained specifically on SIR-based stochastic simulations. They incorporated three types of noise,additive white noise, multiplicative environmental noise, and demographic noise,to simulate the effect of stochasticity in real-world outbreaks. A total of 30,000 time series were simulated across the three noise types, with half containing transcritical bifurcations and half containing null bifurcations. Key parameters, such as disease transmission rates and noise intensity, were randomized during data generation to introduce variability and reflect uncertainty. Miry et al. [3] subsequently extended this with a parallel LSTM-CNN model and demonstrated improved performance. However, these models are not accurate on a state-by-state basis, where there are varying levels of noise and other environmental and/or demographic factors, and a systematic real-world evaluation across U.S. state-level surveillance data has not been conducted.

The present work evaluates these pre-trained models on real-world U.S. influenza hospital admissions data, providing a systematic, state-by-state assessment of their performance.

## 3 Data Preparation

### 3.1 Dataset

The influenza data used in this study are weekly hospital admissions from the CDC FluSight Forecast Hub repository.^1^ Additional CDC hospital respiratory surveillance data were also consulted during the study.^2^ The dataset spans from July 2022 through early 2025, covering all 50 U.S. states and Washington D.C., as well as national-level aggregates. The time series comprises approximately 159 weekly observations per location.

### 3.2 Preprocessing Pipeline

The following steps were applied to prepare the data for input to the deep learning models:

1. Split data by state (and national aggregate); for each location:
2. Add padding at the beginning of the series.
3. Reformat columns to datetime format.
4. Split the data at desired subsections via a windowing function.
5. Convert to an EWSTools TimeSeries object.
6. Detrend the data using a Lowess filter with a span of 0.2, consistent with the preprocessing used in the Bury et al. training pipeline [6].
7. Compute residuals by dividing each value by the mean of the absolute value of the dataset.

For the *R_t_*estimation pipeline (described below), steps 4–7 are omitted; instead, the full raw time series is used.

## 4 Methods

### 4.1 Training Data and Bifurcation Dynamics

The models evaluated in this work were trained on simulated data based on transcritical bifurcations in the SIR model. A disease goes extinct when *R*_0_ < 1 and persists when *R*_0_ *>* 1; a transcritical bifurcation occurs at *R*_0_ = 1, where both equilibria meet and exchange their stability. The basic reproduction number is defined as:

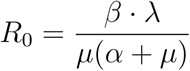

Three types of noise were incorporated to generate stochastic simulations of the SIR model.

#### Additive white noise

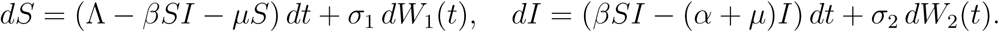

#### Multiplicative environmental noise

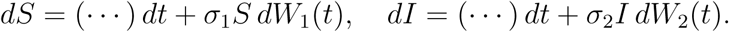

##### Demographic noise

Starting from a discrete event model (birth, infection, recovery, death), the mean and covariance of state changes over Δ*t* are computed. Taking Δ*t →* 0 yields:

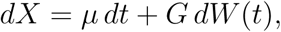

where *G* is derived from the square root of the covariance matrix of the event transitions.

To simulate approaching the transcritical point, the transmission rate *β*_0_ was varied linearly so that *R*_0_ crosses 1:

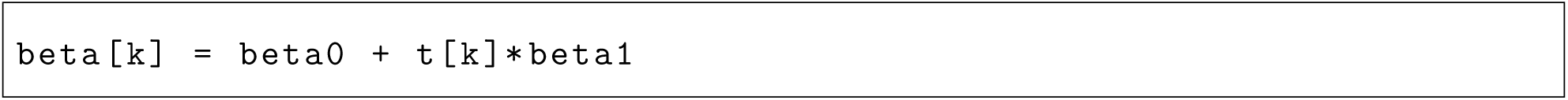

Data were simulated for different parameter values so that some time series occurred below the threshold (no outbreak) and some at or above it (outbreak), allowing the neural network to learn pre-transition versus post-transition patterns.

### 4.2 Estimating the Reproduction Number via EpyEstim

To determine the ground-truth bifurcation point in the real-world data, we estimated the time-varying reproduction number *R_t_*using the Python package EpyEstim, which is based on the EpiEstim framework [4, 8, 5]. The underlying renewal equation methodology originates with Wallinga and Teunis [9]. EpyEstim estimates *R_t_* using a Bayesian renewal equation approach given the mean serial interval (SI) distribution and mean reporting delay distribution.

#### 4.2.1 Probability Mass Function (PMF) Construction

A core requirement of the *R_t_* estimation method is the discretization of continuous probability distributions (Gamma in this case) into a Probability Mass Function (PMF):

**1. Parameterization:** The function accepts a mean (*µ*) and standard deviation (*σ*) and calculates the shape (*α* = (*µ/σ*)^2^) and scale (*β* = *σ*^2^*/µ*) parameters for the Gamma distribution.

**2. Discretization:** The continuous Gamma distribution is discretized using discrete_distrb.

**3. Normalization:** A small floor of 10*^−^*^6^ is applied to all probability values to prevent zero probabilities, and the resulting PMF is normalized so that ^Σ^ *P* (*x*) = 1.

Based on the literature [7], the distributions used were:

1. Serial Interval: *µ_SI_* = 3.6 days, *σ_SI_* = 1.6 days.

2. Reporting Delay: *µ*_Delay_ = 5.8 days, *σ*_Delay_ = 2.8 days.

Since consistent serial interval estimates were unavailable at the state level, *R_t_*estimation was conducted only on national-level data.

#### 4.2.2 Weekly and Daily Incidence Handling

To maintain a weekly incidence resolution, the mean and standard deviation of both distributions are divided by 7 to yield weekly equivalent parameters. Optionally, a synthetic daily series may be reconstructed by evenly dividing each weekly count by seven and interpolating dates:

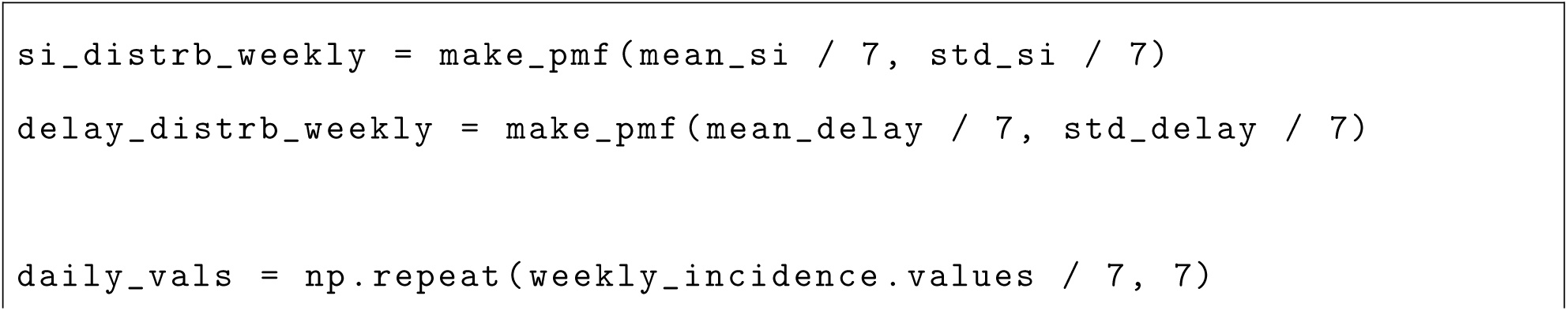
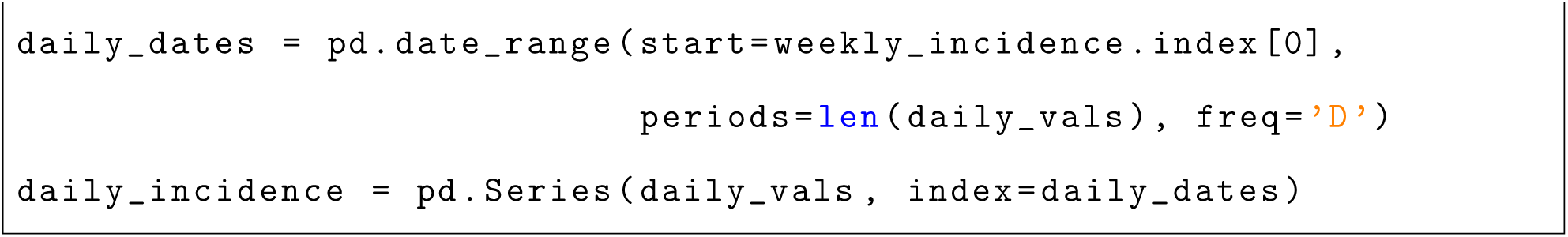

A boolean flag use_daily controls which mode is active: in daily mode, a smoothing window of 7 days is applied to mitigate noise; in weekly mode, the smoothing window is set to 1 as the data are already aggregated.

#### 4.2.3 Rt Estimation

The final *R_t_*estimate is obtained using the bagging_r function:

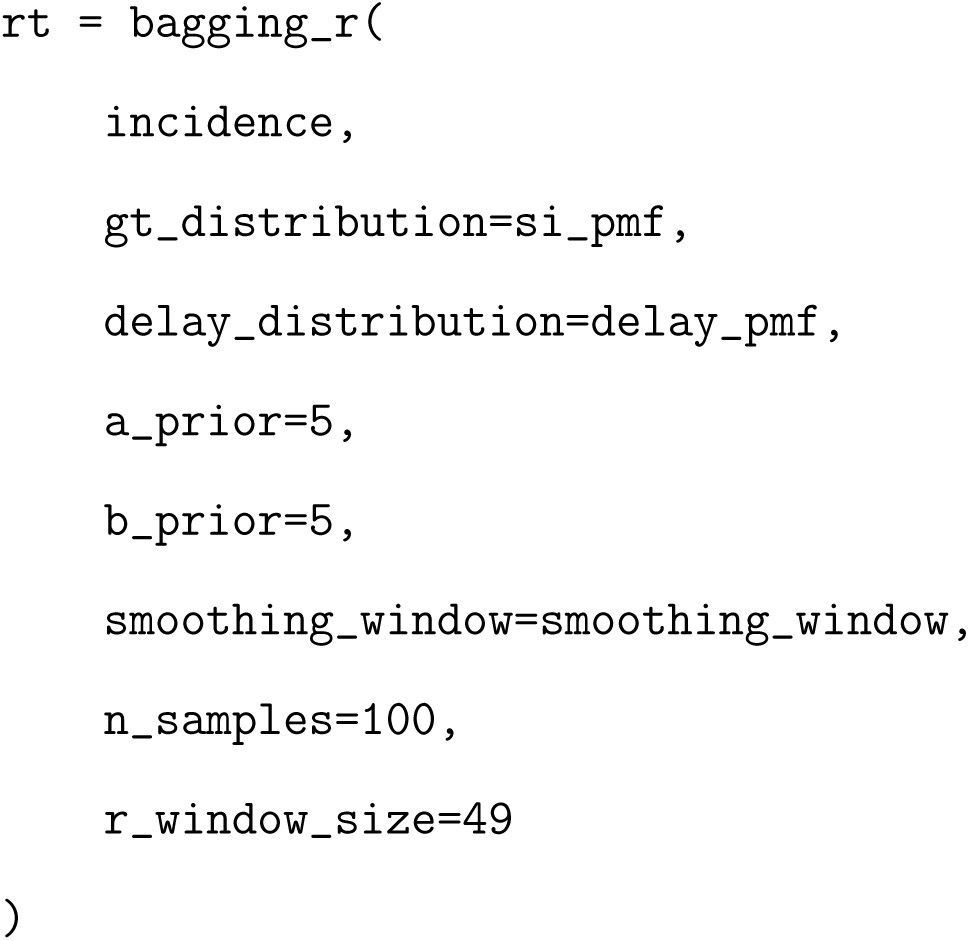

Points in time where the median *R_t_* estimate crosses 1 (from below) were identified as bifurcation points (disease emergence events) and used as ground truth labels for evaluating model performance.

### 4.3 Deep Learning Model: End-to-End Strategy

Before describing our experimental windowing strategies, we detail the end-to-end pipeline of the Bury et al. model [1], covering input requirements, architecture, and how predictions are produced on a time series. The Chakraborty/Miry models [2, 3] follow the same input/output convention and are noted where they differ.

#### 4.3.1 Input: Training Data and Preprocessing

Bury et al. constructed their training set by simulating time series from a large, randomly generated library of ODEs with randomly sampled polynomial terms, producing dynamical systems exhibiting fold, Hopf, and transcritical bifurcations. Two classifier variants were trained: a *500-classifier* trained on 500,000 time series of length 500, and a *1500-classifier* trained on 200,000 time series of length 1,500. The models evaluated in this work accept fixed-length inputs of 100 time steps; shorter series are zero-padded on the left to fill the tensor.

Before being passed to the network—both during training and at inference time—every time series must undergo two preprocessing steps [6]:

**1. Detrending:** The raw series is detrended using Lowess smoothing with a span of 0.2, removing slow trends while preserving the fluctuation structure that encodes critical slowing down.

**2. Normalization:** Each residual series is normalized by dividing every data point by the mean absolute value of the residuals across the full series.

This preprocessing is a strict requirement: because the model was trained exclusively on Lowess-detrended, mean-absolute-value-normalized residuals, deviating from this pipeline at inference time substantially degrades performance [6]. The preprocessing steps in Section 3.2 of this paper mirror this pipeline exactly.

#### 4.3.2 Model Architecture

The CNN-LSTM architecture sandwiches two types of neural network layers. The CNN layer reads in subsequences of the input and extracts local temporal features. The LSTM layer then ingests the CNN output and, through recurrent connections, builds a memory of how those features evolve over time—enabling recognition of the same dynamical pattern at different positions in the series. The network was trained for 1,500 epochs with a learning rate of 0.0005, with hyperparameters tuned via grid search. To reduce variance, ten independently trained models are ensembled by averaging their per-step predictions.

The Chakraborty/Miry models use the same CNN-LSTM arrangement but were retrained on SIR-specific stochastic simulations (Section 4.1). Miry et al. [3] additionally introduced a *parallel* LSTM-CNN variant in which the input passes through LSTM and CNN branches simultaneously before their representations are combined, improving robustness under high-noise conditions.

#### 4.3.3 Output: Predictions on a Time Series

At each time step *t*, the model receives the 100-point preprocessed input tensor and outputs a probability distribution over four classes: fold bifurcation, Hopf bifurcation, transcritical bifurcation, and no bifurcation. For epidemic applications the relevant output is the *transcritical bifurcation probability*, which is thresholded at 0.5 to yield a binary prediction at that time step. Applied sequentially across all *t*, this produces a full probability trace over the time series—one scalar per step—enabling online detection without requiring the bifurcation location to be known in advance.

The key constraint this imposes on real-world deployment is that the 100-point input tensor must be re-populated at every time step. The original Bury et al. code does this by always using the most recent 100 points (zero-padded at the start of the series). How that tensor is constructed as new data arrive is not uniquely prescribed by the original work, and is the subject of the experiments described next.

### 4.4 Windowing Strategies for Online Prediction

Having established the model’s input requirements above, we now describe the windowing strategies we designed and evaluated to assess whether these models are suitable for real-time prediction on real surveillance data. Each strategy defines a different rule for constructing the 100-point input tensor at each time step *t*:

#### Rolling window (last 100 points)

This is the method used in the original Bury et al. code, in which only the last 100 data points are used as input to the model. The window is initially padded with zeros; in each iteration, progressively more of the 100 points are revealed.

#### Showing earlier tipping points

We tested how the model performs when more than one bifurcation is visible within a single input tensor, by feeding windows that span multiple tipping points.

#### Validation cut (points earlier than 60 excluded)

To validate the rolling window results in Section 5.4.1, we reproduced the outputs from that experiment but truncated the non-used data (points earlier than index 60) from each window. This served as a sanity check confirming that the earlier padded region does not materially affect the model outputs.

#### Moving window across entire data

This method shifts a window of fixed length 100 across the entire time series from *t* = 1 to *t* = 159, simulating a sliding-window online detection scenario.

#### Rolling window (length 50, zero-padded)

To reduce the probability that a second bifurcation appears in the input tensor, a shorter rolling window of length 50 is used, with the remaining 50 positions always zero-padded. This trades off the possibility of second-bifurcation contamination against the risk of the model never seeing a complete input.

#### Expanding window (last 100 points)

An expanding window approach is used, in which the window grows from the start of the series up to the current time point, but never exceeds length 100. This retains the model’s full context without exposing it to future data.

### 4.5 Evaluation Metrics

For each windowing strategy, model outputs at each time step were thresholded at a probability of > 0.5 to produce binary predictions. Ground-truth positive labels were assigned to the set of time steps within manually identified bifurcation intervals, and metrics were computed accordingly.

A model was additionally judged “accurate” at the interval level if the mean predicted probability across Interval A (*t* = 108–118) or Interval B (*t* = 127–140) exceeded 60%:

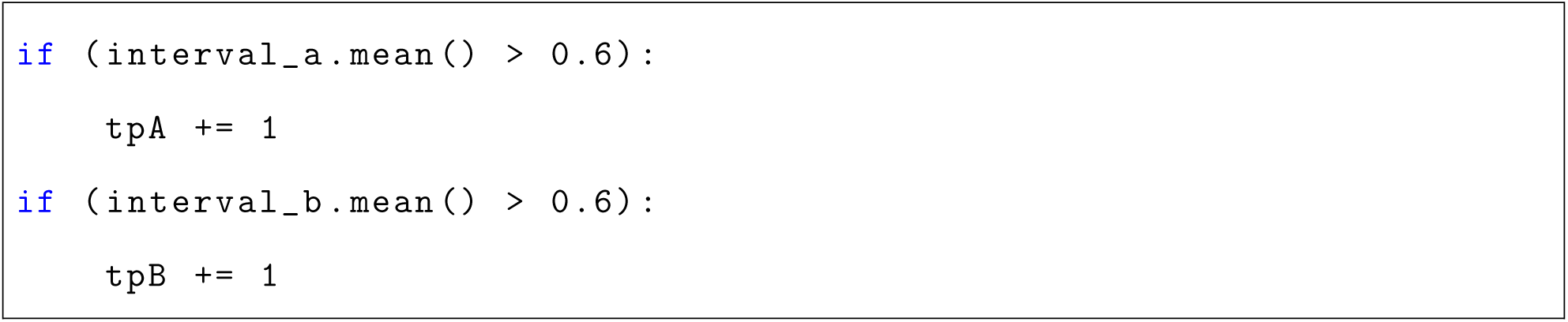

## 5 Results

### 5.1 Reproduction Number Estimation and Ground-Truth Bifurcation Points

Figure 1 shows the national-level weekly hospital admissions time series, which exhibits multiple epidemic waves spanning 2022–2025. Figure 2 shows the estimated *R_t_* alongside the hospital admissions data; the dashed line at *R_t_* = 1 marks the transcritical bifurcation threshold. The *R_t_* trace crosses 1 from below at two distinct epochs corresponding to the onset of major flu waves, which we designate as the two ground-truth bifurcation windows.

**Figure 1:**
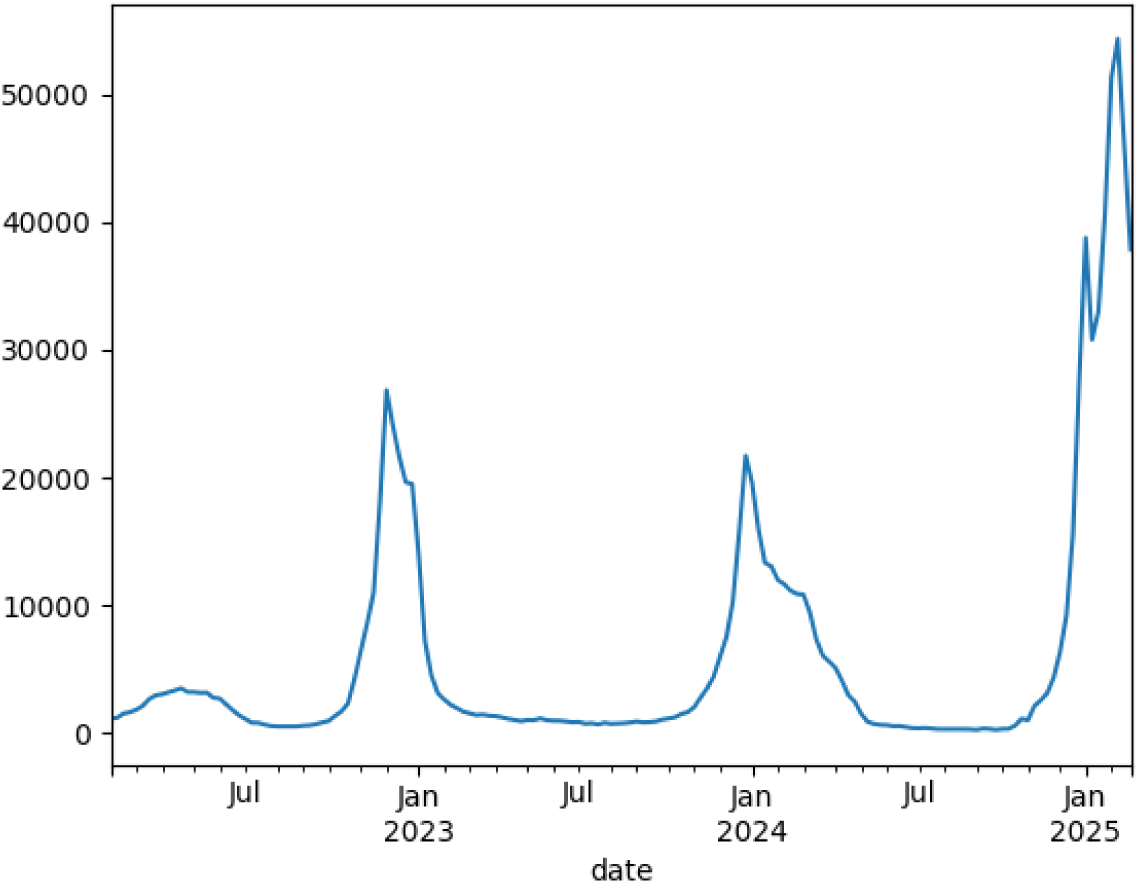
National-level weekly influenza hospital admissions time series, spanning July 2022 to early 2025.

**Figure 2:**
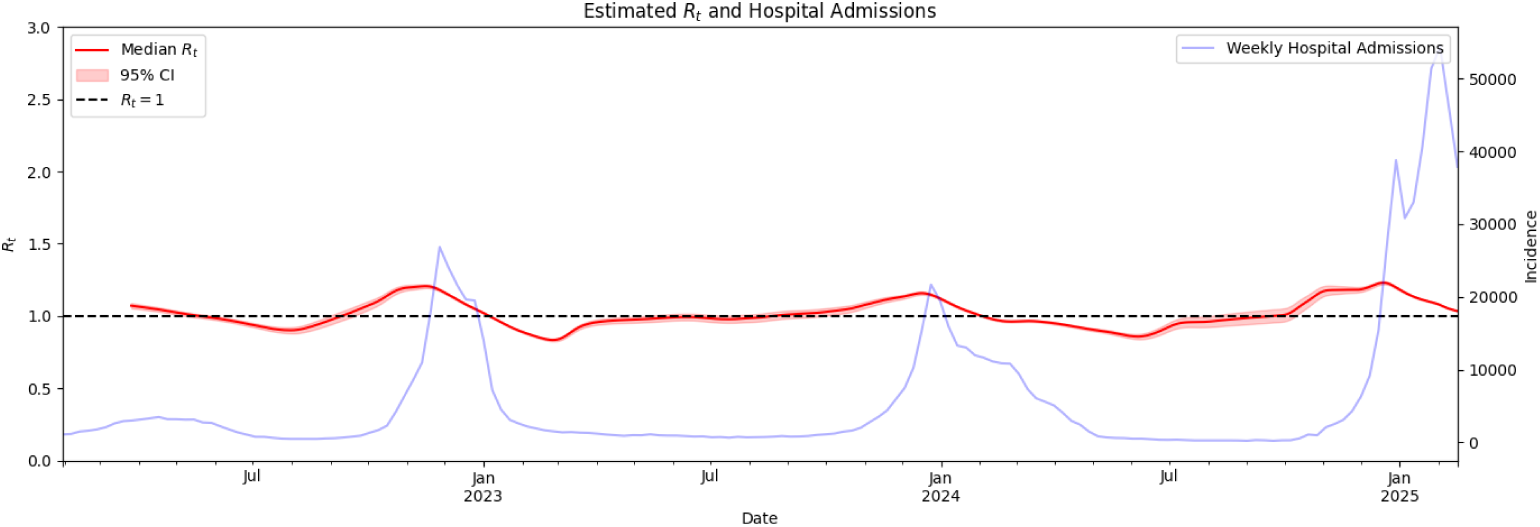
Estimated *R_t_* (median, 95% CI) overlaid with weekly hospital admissions. The dashed horizontal line marks *R_t_* = 1; upward crossings correspond to the identified transcritical bifurcation points.

**Figure 3:**
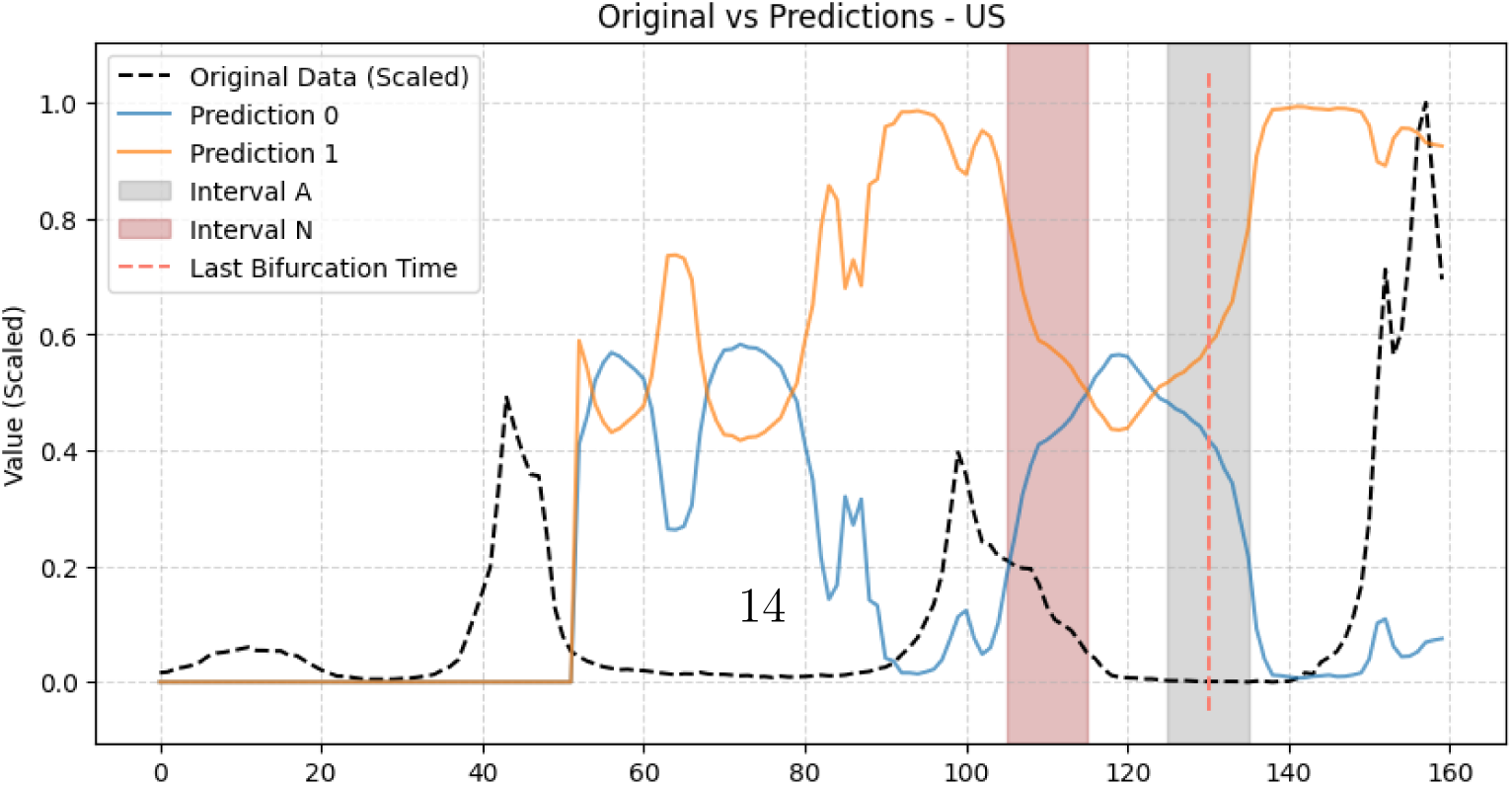
DL bifurcation prediction probabilities on U.S. national data given the estimated real bifurcation point. The model (rolling window, length 100) shows high true positive rate near the identified bifurcation.

### 5.2 State-Level Model Accuracy Across Two Bifurcation Intervals

The models were run on every state. For evaluation, two bifurcation intervals were manually identified: Interval A (*t* = 108–118) corresponds to the first major bifurcation and Interval B (*t* = 127–140) to the second. The chosen intervals were defined as follows:

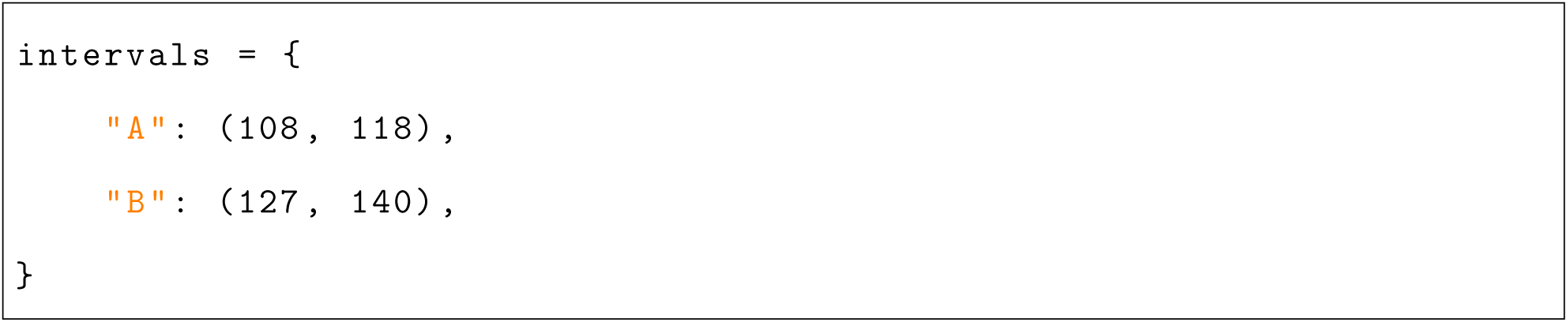

Since the prediction probabilities become more accurate as the window approaches the bifurcation point, it is most informative to evaluate the model within a focused window near the event rather than across the full series. As a result, these intervals were chosen manually to best encompass the peak bifurcation dynamics across all states.

Figure 7 plots each state’s mean DL prediction probability across Interval A (black bars) and Interval B (red bars). The black bars are much more consistently within the range of 90%–98% prediction probability, while the red bars fall in the range of approximately 48%–70%.

**Figure 4:**
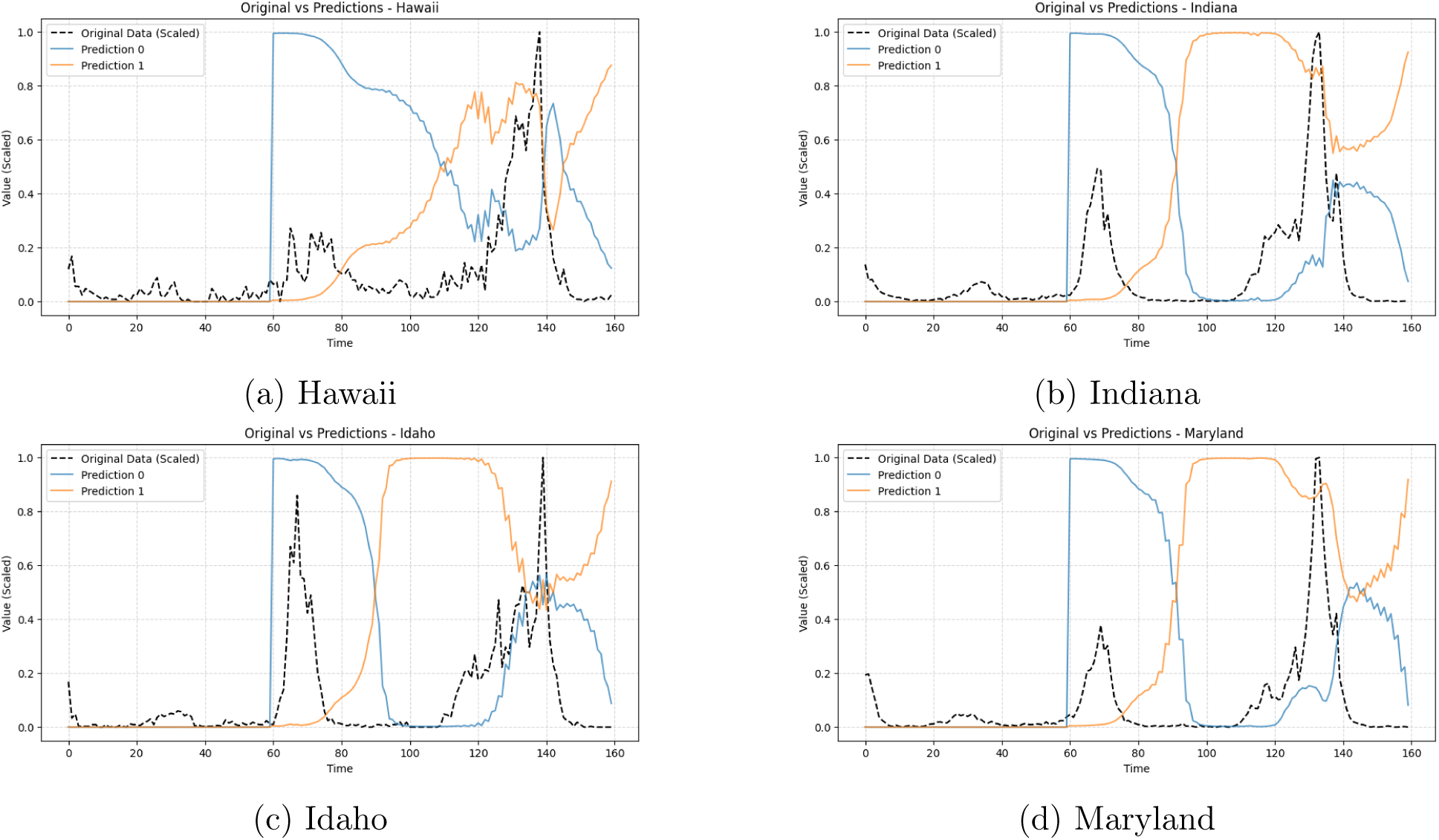
DL bifurcation prediction probabilities and original time series for selected states (Part 1).

**Figure 5:**
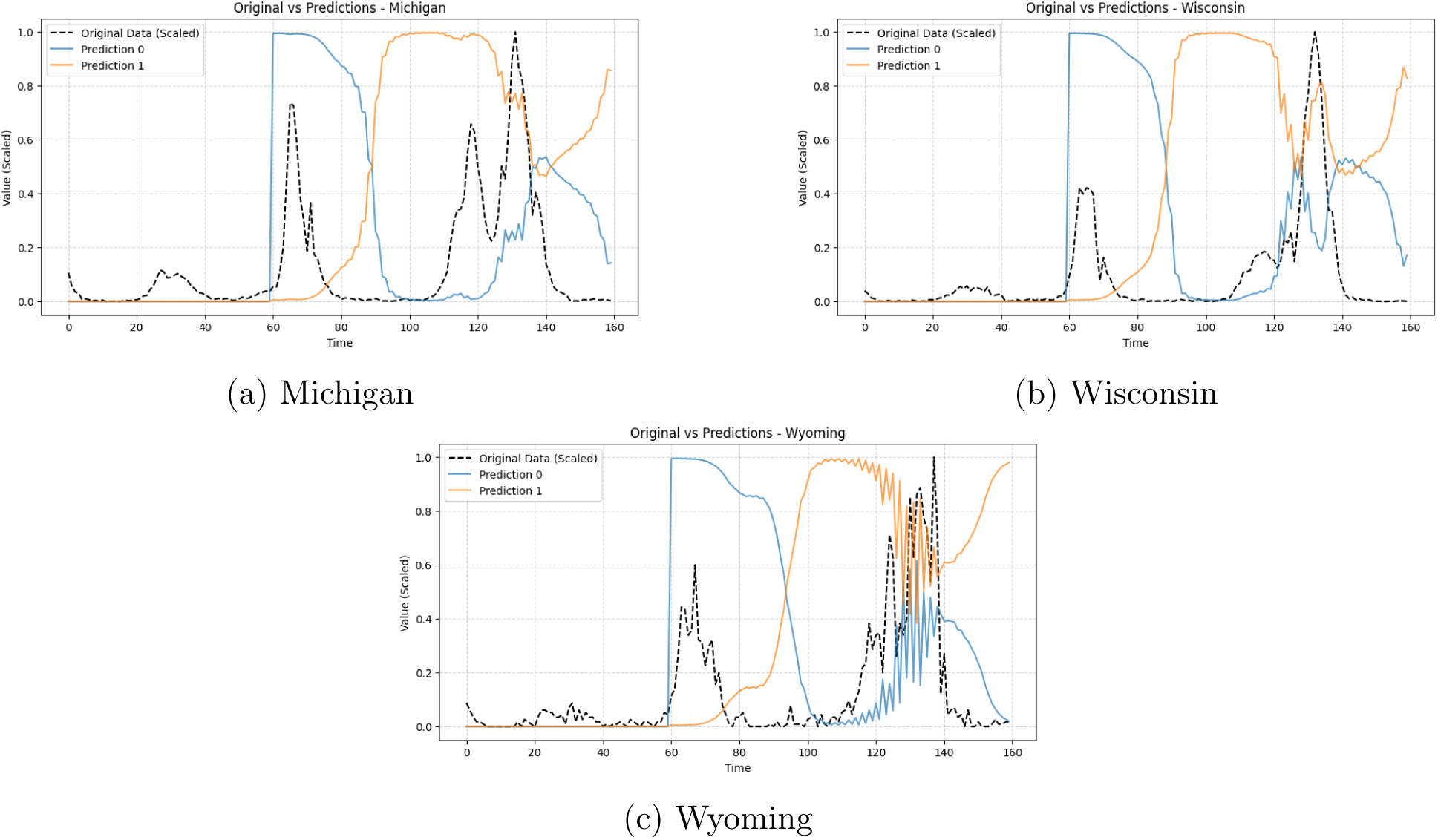
DL bifurcation prediction probabilities and original time series for selected states (Part 2).

**Figure 6:**
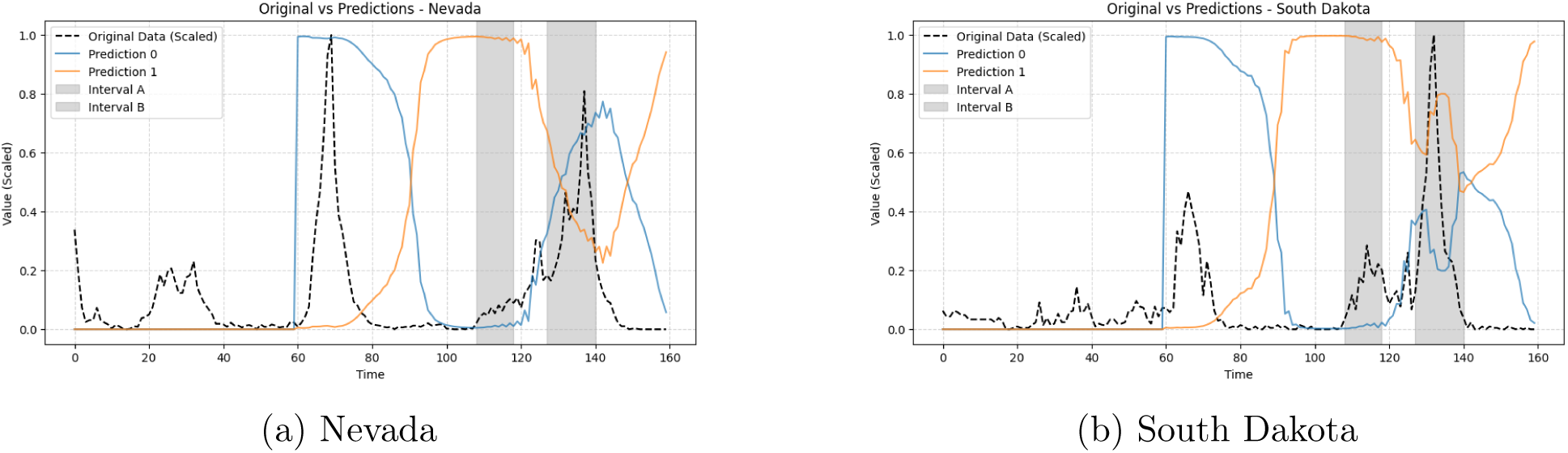
DL bifurcation prediction probabilities and original time series for selected states (Part 3).

**Figure 7:**
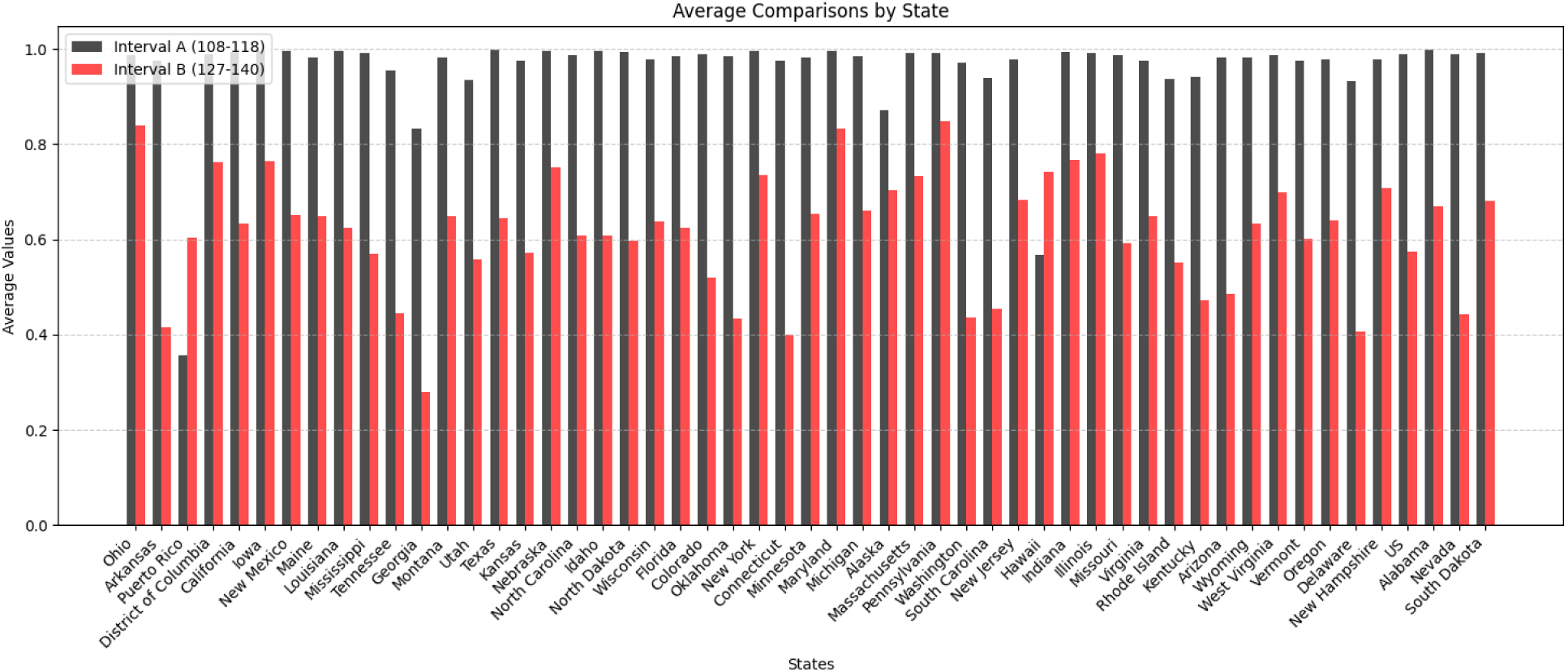
Each state’s mean DL prediction probability across Interval A (108–118) and Interval B (127–140). Black bars correspond to Interval A; red bars to Interval B.

Table 1 summarizes the true positive rates across intervals.

**Table 1:**
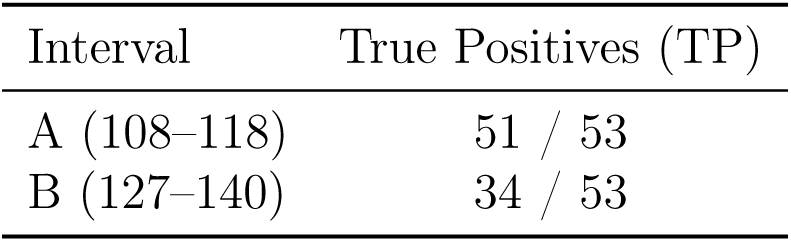
True positive rates by bifurcation interval across all 53 locations (50 states + DC + Puerto Rico + national).

### 5.3 Model Comparison Across Classifiers

Table 2 reports the confusion matrix values for the three pre-trained model variants applied using the rolling 100-point window strategy.

**Table 2:**
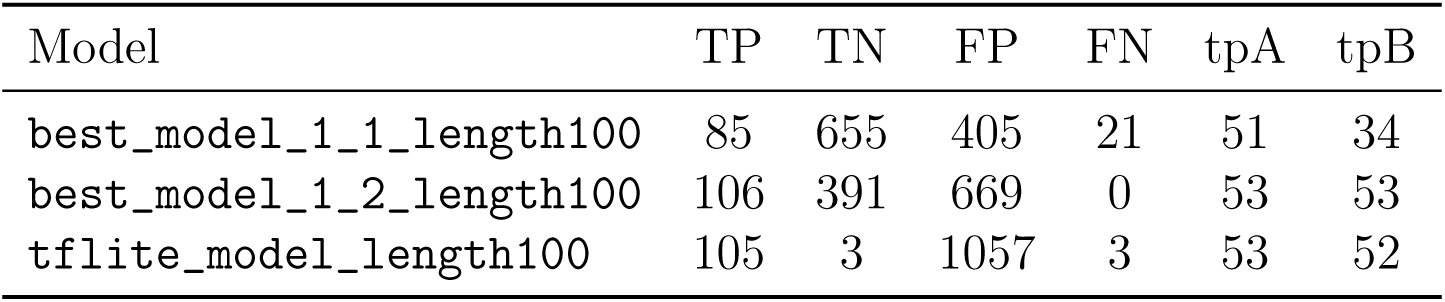
Performance of three pre-trained model variants on all states/locations, rolling window (length 100).

| Model | TP | TN | FP | FN | tpA | tpB |
| --- | --- | --- | --- | --- | --- | --- |
| best_model_1_1_length100 | 85 | 655 | 405 | 21 | 51 | 34 |
| best_model_1_2_length100 | 106 | 391 | 669 | 0 | 53 | 53 |
| tflite_model_length100 | 105 | 3 | 1057 | 3 | 53 | 52 |

### 5.4 Windowing Strategy Experiments

#### 5.4.1 Rolling Window, Last 100 Points (Baseline)

This is the method used in the original code from Bury et al., in which only the last 100 data points are used as input to the model. Initially padded with zeros, in each iteration, progressively more of the 100 points are revealed. Applying this method to best_model_1_1_length100 across all 53 locations yielded:

• TP: 85, TN: 655, FP: 405, FN: 21

• Accuracy: 63.48%, Precision: 17.35%, Recall: 80.19%, F1 Score: 28.52%

#### 5.4.2 Showing Earlier Tipping Points in the Input Tensor

We tested how the model performs when more than one bifurcation is visible within a single input tensor,specifically, feeding windows that span both bifurcation epochs simultaneously near Interval A (*t* = 108–118). The raw confusion matrix counts from this experiment are as follows:

• tpA: 46, tpB: 20

• TP: 66, FP: 507, TN: 593, FN: 44

• Accuracy: 54.46%, Precision: 11.52%, Recall: 60.00%, F1 Score: 19.33%

Visually, this confirms that showing more than one tipping point in a single input tensor can decrease accuracy for each individual bifurcation interval. However, as discussed below, aggregate metrics may behave differently depending on how the evaluation windows are defined.

#### 5.4.3 Validation: Cutting Off Points Earlier than Index 60

To confirm the baseline rolling window results (Section 5.4.1), we reproduced those outputs with the non-active data (indices earlier than 60) removed from each input window. This served purely as a validation step, confirming that the zero-padded prefix region of the tensor does not materially influence model outputs.

#### 5.4.4 Moving Window Across Entire Data

This method shifts a window of fixed length 100 across the entire time series, starting at *t* = 1 and ending at *t* = 159, simulating a sliding-window online detection scenario. Applying this method across all states:

• tpA: 38, tpB: 43

• TP: 81, FP: 259, TN: 841, FN: 29

• Accuracy: 76.12%, Precision: 23.82%, Recall: 73.64%, F1 Score: 35.96%

Note that in this configuration multiple tipping points were still sometimes visible within the same input tensor; despite this, overall accuracy increased relative to the baseline, supporting the conclusion that sufficient coverage of the first tipping point is more important than excluding subsequent ones.

#### 5.4.5 Rolling Window, Length 50 (Zero-Padded)

To reduce the probability that a second bifurcation appears in the input tensor, a shorter rolling window of length 50 was applied, with the remaining 50 positions always zero-padded. This method yielded:

**• Accuracy: 79.72%, Precision: 55.56%, Recall: 94.34%, F1 Score: 69.93%, Specificity: 74.84%**

While this strategy mitigates second-bifurcation contamination, the model never receives a fully populated 100-point input, which can inhibit performance in other ways.

#### 5.4.6 Expanding Window, Last 100 Points

An expanding window approach was used, in which the window grows from the start of the series up to the current time point but never exceeds length 100. This retains the model’s full temporal context from the start of the series and yielded the best overall performance:

• **Accuracy: 90.09%, Precision: 72.86%, Recall: 96.23%, F1 Score: 82.93%, Specificity: 88.05%**

### 5.5 Windowing Strategy Summary

Table 3 summarizes performance metrics across all windowing strategies applied to best_model_1_1_lengt

**Table 3:**
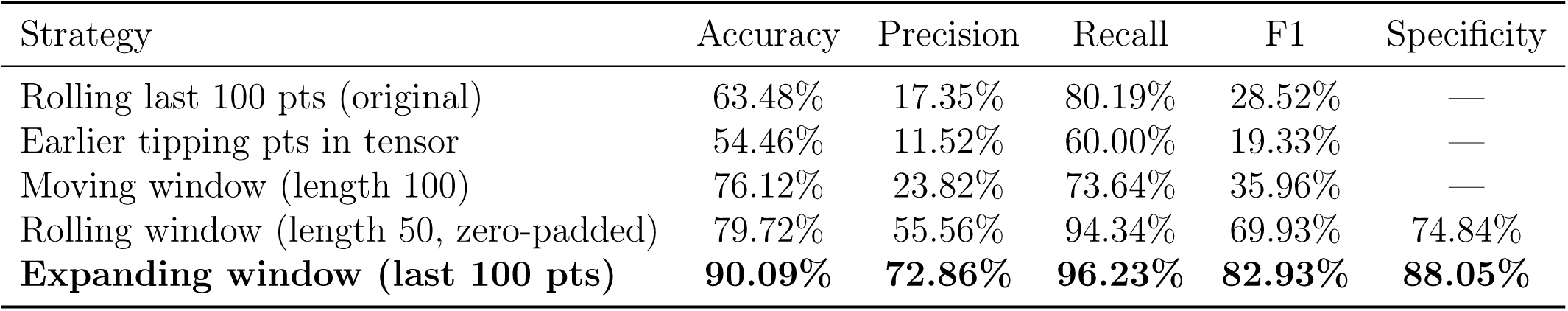
Performance metrics across windowing strategies.

Key observations from the windowing experiments:

- Showing more than one tipping point in a single input tensor *can* decrease accuracy visually for each individual interval, but aggregate metrics may still improve because the model sees more bifurcation-related data.
- When applying the model in real-time (simulated by moving the input tensor throughout the time series), performance generally increases or stays the same.
- The expanding window approach, which retains all historical data up to the current time (bounded at 100), achieves the best overall metrics.
- As long as a sufficient portion of the first tipping point is included in the input tensor, model accuracy will not be hindered and could potentially even be improved.

## 6 Discussion

### 6.1 Key Findings

Using the methodologies described above, and with visual analysis, it does seem that showing more than one tipping point in an input tensor can decrease accuracy for individual intervals. However, looking at the accuracy metrics in aggregate, the model performs better when it receives more bifurcation-containing data. Furthermore, when applying this in real-time (simulated by moving the input tensor throughout the time series), performance does seem to increase or stay the same. This points to a conclusion that as long as a sufficient portion of the first tipping point is included, the model accuracy will not be hindered, and could possibly even be improved. Thus, from this analysis, this model is capable of application to real-world data and online live predictions.

The expanding window approach yields the strongest overall performance (accuracy 90.09%, F1 82.93%), suggesting that providing the model with the full temporal context from the start of the series is beneficial.

### 6.2 Accuracy Metrics and the Two Bifurcation Intervals

These accuracy metrics were computed by including two different types of bifurcations. Given the pre-trained models’ input data, the models always perform well on Interval A bifurcations, while performing lower on Interval B bifurcations,this is the main reason why the overall accuracy metrics are lower than expected.

It seems that the model is pretty consistent for the first major bifurcation (Interval A); however, there does seem to be variance in the second bifurcation. This could be because it is hard to choose an interval that encompasses all peak bifurcations. As for each state, the bifurcation timing in Interval B varies between states, so it is hard to choose a time frame (without it being too large) that can encompass all of them. The model is much more accurate for bifurcations that more accurately resemble the bifurcations in the training data. This sounds trivial,the model was not trained on bifurcations similar to those found in Interval B, which explains the drop in prediction accuracies.

### 6.3 Limitations

The *R_t_*-based ground truth bifurcation points could only be computed at the national level, since consistent serial interval values for U.S. state-level data were unavailable. For state-level evaluation, manually chosen interval windows were used instead. Additionally, the precision values across most windowing strategies are relatively low, driven by a large number of false positives in the non-bifurcation portions of the time series,suggesting these models may benefit from threshold tuning for operational deployment.

### 6.4 Markov Regime Switching as a Possible Alternative

One possible alternative to the deep learning approach is the application of Markov Regime Switching (MRS) models for detecting bifurcations. MRS models assume that a time series can be described by several submodels (states or regimes), where switching between submodels is governed by a hidden Markov chain. In the epidemic context, these states can correspond to endemic and outbreak phases. We discuss MRS models as a promising complement to the DL approach, though a full comparative evaluation is left to future work.

## Data Availability

All data used in this study are publicly available from the U.S. Centers for Disease Control and Prevention (CDC) via the FluSight Forecast Hub and data.cdc.gov (links provided below). All code used for data preprocessing, reproduction number estimation, and model evaluation is openly available on GitHub at the repositories listed below.

https://raw.githubusercontent.com/cdcepi/FluSight-forecast-hub/09a9d11e52413297732c83d36bcdd8e1a0a13fc9/target-data/target-hospital-admissions.csv

https://data.cdc.gov/Public-Health-Surveillance/Weekly-Hospital-Respiratory-Data-HRD-Metrics-by-Ju/ua7e-t2fy/about_data

https://data.cdc.gov/Public-Health-Surveillance/Weekly-Hospital-Respiratory-Data-HRD-Metrics-by-Ju/mpgq-jmmr/about_data

https://github.com/burakayy7/deep_learning_bifurcation_prediction_collection_V1

https://github.com/burakayy7/fluBifurcation_deep_learning

https://github.com/burakayy7/dl_ews_training

https://github.com/burakayy7/bifurcationDetection_deep_learning

https://github.com/burakayy7/ewstools-flu-dataset

## Code Availability

All code developed for this study is available at the following repositories:

- https://github.com/burakayy7/deep_learning_bifurcation_prediction_collection_V1/tree/main
- https://github.com/burakayy7/fluBifurcation_deep_learning
- https://github.com/burakayy7/dl_ews_training
- https://github.com/burakayy7/bifurcationDetection_deep_learning
- https://github.com/burakayy7/ewstools-flu-dataset

## Acknowledgments

The authors thank the UVA Biocomplexity Institute for support and resources.

## Footnotes

1 https://raw.githubusercontent.com/cdcepi/FluSight-forecast-hub/09a9d11e52413297732c83d36bcdd8e1a0a13fc9/target-data/target-hospital-admissions.csv

2 Weekly Hospital Respiratory Data by Jurisdiction: https://data.cdc.gov/Public-Health-Surveillance/Weekly-Hospital-Respiratory-Data-HRD-Metrics-by-Ju/ua7e-t2fy/about_data; by State: https://data.cdc.gov/Public-Health-Surveillance/Weekly-Hospital-Respiratory-Data-HRD-Metrics-by-Ju/mpgq-jmmr/about_data

